# Intracerebral hemorrhage induces monocyte TNF signaling that is suppressed by Siponimod (BAF312): a single-cell transcriptomics study in patients

**DOI:** 10.64898/2026.01.22.26344292

**Authors:** Jonathan Howard DeLong, Sebastian Díaz-Pérez, Kevin Navin Sheth, Jang-Ho Cha, CJ Malanga, Patricia GM Wagner, Nicole Pezous, Aurelie Hanin, Kyle B. Walsh, H.E. Hinson, Lauren Hachmann Sansing

## Abstract

Intracerebral hemorrhage (ICH) causes high morbidity and mortality, with neurotoxic inflammation driven by infiltrating monocytes. Therapeutic options remain limited. Here we performed single-cell RNA sequencing and plasma cytokine analysis on peripheral blood samples from ICH patients treated with the immunomodulatory drug BAF312 (Siponimod) or placebo at days 1, 3, and 7 post ICH. In the absence of treatment, the inflammatory response peaked at day 3 post ICH. BAF312 markedly reduced peripheral blood T and B lymphocyte numbers by day 3. BAF312 also impacted the myeloid response, suppressing TNF signaling in classical and non-classical monocytes. Multiple cytokine signaling pathways were decreased, though BAF312 did not impact plasma cytokine or chemokine concentrations. Notably, increased monocyte TNF signaling correlated with better functional outcome, possibly related to the positive role of monocytes during the subacute stage of ICH. These findings suggest that BAF312 suppresses peripheral immune responses after ICH and supports a complex role of monocytes in this disease.

## INTRODUCTION

Intracerebral hemorrhage (ICH) is the second most common form of stroke, affecting 2 million people annually worldwide [1, 2]. It is the deadliest type of stroke, with a 30-day mortality rate of roughly 40% [3-5]. The influx of blood into the brain parenchyma initially damages the brain by mass effect, but this is rapidly followed by cascades of secondary injury. Neurons die through both necroptotic and ferroptotic pathways [6]. Molecules released from these inflammatory cell death pathways, as well as from blood components, activate microglia, astrocytes, and endothelial cells. This activation leads to the recruitment of blood monocytes to the brain, which differentiate into macrophages that contribute to inflammatory damage in the acute and early subacute stages of ICH [7]. In addition, T cells infiltrate the brain by day 1 in murine models of ICH, weakening the blood-brain barrier and worsening functional outcome [8, 9]. After several days, macrophages are found to primarily contribute to reparative processes including phagocytosis of cell debris [7]. Regulatory T cells may also play a role in limiting inflammation, BBB breakdown, and neurological deficits after ICH [10, 11]. Thus, there is significant interest in the developing therapeutics that can limit inflammation-dependent damage in the acute stage of ICH without impairing the reparative roles of immune cells in subsequent stages.

BAF312 (Siponimod) is a small molecule modulator of sphingosine-1-phosphate (S1P) receptors. S1P is a bioactive lipid secreted by activated platelets, erythrocytes, and endothelial cells, and signals through five S1P receptors (S1PR1-S1PR5), which are highly expressed by endothelial cells, platelets, monocytes, lymphocytes, microglia, neurons, astrocytes, and oligodendrocytes. BAF312 selectively binds S1PR1 and S1PR5, downregulating S1PR1 expression and acting as a functional S1PR1 antagonist [12]. Developed as an improved alternative to the S1P analog FTY720 (fingolimod), BAF312 has more selective binding and a reduced elimination half-life [12-14]. BAF312 can cross the blood-brain barrier (BBB) and in contrast to FTY720 does not need to be phosphorylated *in vivo* to target S1P receptors. BAF312 and FTY720 are approved for treatment of multiple sclerosis, with efficacy thought to depend at least in part on sequestration of T cells and B cells in secondary lymphoid organs [12, 15-19].

In addition to their effects on lymphocytes, BAF312 and FTY720 also act on ICH-relevant non-lymphocyte cell populations relevant to ICH [20]. For example, BAF312 limits transmigration of human PBMC across an endothelial monolayer *in vitro* by an astrocyte-dependent mechanism involving suppression of CCL2 and ICAM-1 expression [21]. Similarly, FTY720 limits production of CCL2 by human U373 astrocytoma cells *in vitro* and reduces astrogliosis in a mouse photothrombotic model of stroke [22, 23]. In murine microglia, FTY720 reduces production of TNF, IL-1β, and IL-6, while increasing production of the neurotrophic factors BDNF and GDNF *in vitro* [24]. These findings suggest that S1P receptor modulators may influence multiple cell types involved in the inflammatory response after ICH.

BAF312 and FTY720 have been shown to reduce inflammation and improve outcome in rodents up to 14 days post ICH or subarachnoid hemorrhage [25-30]. A clinical trial also found that FTY720 decreases neurological impairment in humans during ICH, though the mechanism remains poorly understood [31, 32]. FTY720 and BAF312 are associated with decreased brain edema, fewer brain-infiltrating T cells, less white matter loss, and modest improvements in behavioral outcomes. The effect sizes in these studies tend to be small and effects are not seen in all studies [33], but there are no approved treatments for ICH and even modest effects in preclinical models warrant further investigation in humans.

Despite several clinical trials testing surgical and medical interventions during ICH, there are still no approved therapies available to patients and there remains a compelling need to continue testing novel therapeutics [34]. Additionally, patient samples gathered during clinical trials have been proven to be a valuable source of insight into the basic biology of the disease, even when the therapeutic was not demonstrated to significantly improve patient outcome [32, 35, 36]. RNA sequencing is a powerful tool to characterize transcriptomic profiles in an unbiased manner. Single-cell RNA sequencing allows a researcher to examine cell subsets at higher resolution without introducing a sorting step that might bias the sample or exclude populations of interest. Multiple studies have used RNA sequencing to gain valuable insights into the biology of intracerebral hemorrhage [35-42]. However, extant studies overwhelmingly examine animal models of ICH and/or utilize bulk RNA-sequencing of whole peripheral blood, with limited ability to distinguish the roles of different cell lineages. The field would benefit from transcriptome-wide analysis of human samples at the single-cell level.

The present study examined peripheral blood from 8 patients who suffered an intracerebral hemorrhage and were treated with BAF312 or placebo. We found that inflammatory responses peaked at day 3 in the peripheral blood of placebo-treated patients, mirroring the time-course previously reported in the hematoma [35]. Analysis of 3 published datasets [37-39] examining bulk PBMC from a total of 190 humans and 10 pigs revealed that the genes upregulated after ICH in PBMC were highly expressed by monocytes in our samples. This validated the gene expression seen in our samples and underscored the importance of peripheral monocytes during ICH. In BAF312-treated patients, peripheral blood monocytes exhibited reduced responsiveness to multiple cytokines by day 3 of treatment, suggesting that BAF312 limits innate inflammatory responses even at early timepoints after treatment. Notably, and contrary to expectations, patients whose monocytes exhibited increased TNF signaling had better functional outcomes, suggesting that the beneficial roles of active monocytes during ICH may outweigh the potentially negative direct effects of TNF [43-46].

## METHODS

### Study design

Patient enrollment was performed at 3 approved study sites in the United States according to the parent Stage 2 clinical trial to study the Efficacy, Safety, and Tolerability of BAF312 in patients with ICH, performed by Novartis Pharmaceuticals (ClinicalTrials.gov ID: NCT03338998). This was a randomized, placebo-controlled, subject- and investigator-blinded study of subjects 18-85 years of age with an acute supratentorial ICH 10-60 mL within 24 hours of enrollment. Additional trial details can be found at https://clinicaltrials.gov/study/NCT03338998.

BAF312 was administered daily by intravenous up titration on days 1 to 7 and then by 10mg tablets taken orally on days 8 to 14. Participants were followed to day 90 for neurological (mRS and NIHSS) and safety conditions during 3 clinic visits. Recruitment for the trial was put on hold due to the COVID-19 pandemic. Thirty-two patients were enrolled in the trial and completed the protocol as planned. After 7 months of the trial being on hold, an Interim analysis was conducted and reviewed by the Data Monitoring Committee and the trial was terminated due to lack of potential clinical efficacy. Prior to the trial being put on hold, peripheral blood samples from 8 patients were collected in ACD tubes (BD 364606) at approximately 1, 3, and 7 days post ICH and were shipped to our lab overnight in NanoCool refrigerated shippers (Path-Tec, Midland, GA), consistent with the methods we previously described to preserve transcriptional profiles.[47]

### Sex as a biological variable

Previous studies have found increased type I interferon responses in females during sterile neuroinflammation including intracerebral hemorrhage [48]. This is a substudy of a larger clinical trial and due to a limited number of patients, every sample that became available was collected and used. Both males and females were represented, but 7 of the 8 patients assessed were female, which precludes meaningful quantification of sex effects in this study.

### Collection of patient clinical data

CT scans were performed at approximately day 1, day 8, and day 15 post ICH. ICH volume, perihematomal edema volume, and relative perihematomal edema volume were determined by CT scan. Functional outcome testing was performed by trained, blinded investigators at each study site.

### Peripheral blood sample processing

Peripheral blood mononuclear cells were isolated by Ficoll gradient centrifugation as previously described [35] and were then counted and cryopreserved. All 3 timepoints for a given patient were thawed and analyzed by 10X on the same day to limit longitudinal batch effects. Live singlet cells were sorted on a BD FACSAria. Samples from patients 3, 4, 6, 7, and 8 were stained with BioLegend TotalSeq-A or TotalSeq-B antibodies to allow for multiplexing. Samples from patients 3 and 4 were thawed and analyzed together, as were samples from patients 7 and 8. The cells were then counted and analyzed on the 10X Genomics platform. Cells from 1 patient were found to be of low viability after thawing and were not analyzed by RNA sequencing. Plasma from this patient was included in cytokine measurements by CBA. In total, samples from 7 patients were analyzed by single-cell RNA-sequencing, of which 2 were treated with placebo and 5 were treated with BAF312. One of the patients receiving BAF312 experienced bradycardia after 2 days of treatment and then discontinued treatment. This patient is excluded from all analyses except the correlation of gene expression with clinical characteristics and outcome shown in Fig 5.

### Flow cytometry

Cells were analyzed on a BD FACSAria while sorting for total live singlets. Cells were stained with CD45 PE-Cy7 (BioLegend 304016), CD14 PE (BioLegend 301806), CD56 FITC (BD 562794), CD3 APC-eFluor 780 (eBioscience 47-0036-42), CD4 violetFluor 450 (BD 560345), and CD8 V500 (BD 560774), as well as LIVE/DEAD Fixable Red Dead Cell Stain (Thermo L34972). Data was analyzed using FlowJo v10.10.0 (BD).

### Single cell RNA-sequencing data processing

Fastq files were aligned to the GRCh38-2020-A genome assembly available from 10X genomics using Cell Ranger count v7.1.0. Cell Ranger sequencing quality metrics can be found in Supp Table 1. Aligned reads were processed in R version 4.3.2 using the standard pipeline of Seurat version 4.3.0. RNA counts from all patients and timepoints were normalized using SCTransform and were integrated using reciprocal PCA reduction. The counts data were clustered at high resolution to identify all captured cell populations. Multiplexed samples were demultiplexed using HTODemux() according to the Seurat pipeline. Cells with fewer than 500 RNA features, more than 6000 RNA features, or more than 25% mitochondrial genes were excluded. After quality control filtering, 103,301 total cells were analyzed from the 21 samples (3 timepoints from each of 7 patients). Cluster annotation was performed manually with the aid of SingleR [49] version 2.2.0 to map datasets from Mulder et al 2021 (for monocyte and DC populations) [50] and Hao et al 2021 (for all populations) [51] onto our dataset. Clusters were pooled into a total of 26 populations and doublets and dead cells were removed (Supp 1A, B). These populations were further pooled into 7 lineages for downstream analysis. Proportions of cells in each lineage are shown for each patient in Supp Fig 1C. To limit the disproportionate impact of samples in which more cells were recovered, each population was downsampled to a number roughly proportional to the average number of cells recovered for that lineage. CD16^+^ monocytes, NK cells, CD8^+^ T cells, and B cells were downsampled to 400 cells, CD4^+^ T cells were downsampled to 800 cells, and classical monocytes were downsampled to 1600 cells (Supp 1D).

Differential gene expression was performed using the FindMarkers() function, utilizing the Wilcoxon Rank Sum test with a log fold change threshold of 0.3.

Module scoring was performed using the AddModuleScore() function of Seurat. Scoring of TNF signaling used genes of the MSigDB HALLMARK_TNFA_SIGNALING_VIA_NFKB geneset. Scoring of genes from Bai 2024 used genes that were upregulated or downregulated in bulk RNA sequencing of peripheral blood by at least 2-fold in ICH patients compared to individuals with hypertension. Scoring of genes from Durocher 2019 used genes that were upregulated or downregulated in bulk RNA sequencing of peripheral blood by at least 1.6-fold in ICH patients compared to individuals with matched vascular risk factors. Scoring of genes from Walsh 2019 used the top 100 DEGs found by bulk RNA sequencing of peripheral blood of pigs 6 hours post-ICH compared with blood drawn before ICH induction.

To simplify the analysis of associations between disease severity and gene expression, each patient was placed into one of two groups for each clinical measurement. Good outcome was defined as an mRS below 4, as these patients have moderate disability and are able to walk without assistance. Other metrics were dichotomized at thresholds that would divide the patient population into two roughly equal groups, while avoiding thresholds that would split patients with very similar measurements into opposing groups.

For some analyses examining the association of BAF312 or patient outcome with peripheral immune cell gene expression, cells were pseudobulked within each tissue sample by cell lineage. This aggregated gene counts for all cells within each cell lineage, within each tissue sample, as if each of these populations had been analyzed by bulk RNA sequencing. This further corrected for cell number differences between samples[52] and enabled assessment of differential gene expression using a linear regression model that included a covariate for ICH score, which controlled for baseline differences between our BAF312-treated and placebo-treated patients. Differential gene expression analysis for pseudobulked data was performed using the glmQLFTest() function in the edgeR library.

Ingenuity Pathways Analysis Upstream Regulators analysis was performed using a Benjamini-Hochberg adjusted p-value cutoff of 0.05. Bias-corrected activation z-scores are shown.

Geneset enrichment analysis (GSEA) was performed with the human MSigDB Hallmark Gene Sets using the fgsea v1.28.0 library in R. Single-cell DEG lists were ranked by logFC and pseudobulked DEG lists were ranked by t-statistic calculated with the eBayes() function in the limma v3.58.1 library.

### Cytometric Bead Array

Concentrations of cytokines and chemokines in plasma isolated from patient peripheral blood samples were determined via Cytometric Bead Array (CBA) (BD Biosciences), according to the manufacturer’s protocol. The following CBA Flex Sets were used: TNF (558273), IL-1α (560153), IL-1β (558279), IL-6 (558276), CCL2 (558287), CXCL10 (558280), CD40L (560305), IFNγ (558269), IL-10 (558274), and IL-4 (558272). Concentration measurements that fell below the lower limit of detection (LLOD) were substituted with values equal to half of the lower LLOD (LLOD / 2).

### Statistics

Differential gene expression for single cells was calculated using the FindMarkers() function of Seurat in R, utilizing the Wilcoxon Rank sum test, with a log fold change threshold of 0.3 and excluding genes expressed by less than 10% of cells. P-values were adjusted for multiple comparisons using the Bonferroni correction. Differential gene expression for pseudobulked data was performed by fitting the data to a negative binomial generalized linear model using the glmQLFit() and glmQLTest() methods of edgeR in R. DEGs were extracted using the topTags() function of edgeR, with p-values adjusted by the Benjamini-Hochberg method. All differentially expressed genes can be found in supplemental data, with no p-value or log fold-change cutoffs.

GSEA was performed using the fgsea() function of the fgsea package in R. Shown p-values are adjusted for multiple comparisons using the Benjamini-Hochberg method. Adjusted p-values of GSEA pathways less than 0.05 were considered significant.

Ingenuity Pathways Analysis Upstream Regulators Analysis was calculated using DEGs with adjusted p-values below 0.05. IPA pathways containing fewer than 4 DEGs or with Benjamini-Hochberg-corrected p-values above 0.05 were excluded.

Statistics for cytokine concentrations were determined using the rstatix package in R: p-values were determined by the Wilcoxon Rank Sum Test using the wilcoxon_test() function and corrections for multiple testing was done by Holm’s method using the adjust_pvalue() function. Adjusted p-values less than 0.05 were considered significant. In box and whisker plots, the lower and upper hinges correspond to the first and third quartiles. The whiskers extend to the highest or lowest values no further than 1.5 * the inter-quartile range from the upper or lower hinge, respectively.

### Study approvals

The Human Investigations Committee (IRB) of Yale University approved this study. Patients or their designated surrogates provided written informed consent according to the Declaration of Helsinki.

### Data availability

Values for all data points in graphs are reported in Supp Table 11 - Supporting Data Values. Raw and processed RNA-sequencing data are available in PubMed Gene Expression Omnibus (GEO) database under the accession number GSE313043.

## RESULTS

### Patient baseline characteristics

Peripheral blood samples from 8 patients were collected at approximately 1, 3, and 7 days post intracerebral hemorrhage (Fig 1A). Patient mean age was 65±12 years and 7 of the 8 patients were female (Table 1). Baseline ICH volume ranged from 7.3-48.5 mL, with a mean of 27.9±14.4 mL. Four hemorrhages were located in the basal ganglia and 3 in the occipital lobe. Baseline NIHSS ranged from 3-19, with a mean of 11.5±6.2. Additional patient demographics, clinical characteristics, and outcome data can be found in Supp Table 2.

**Figure 1.**
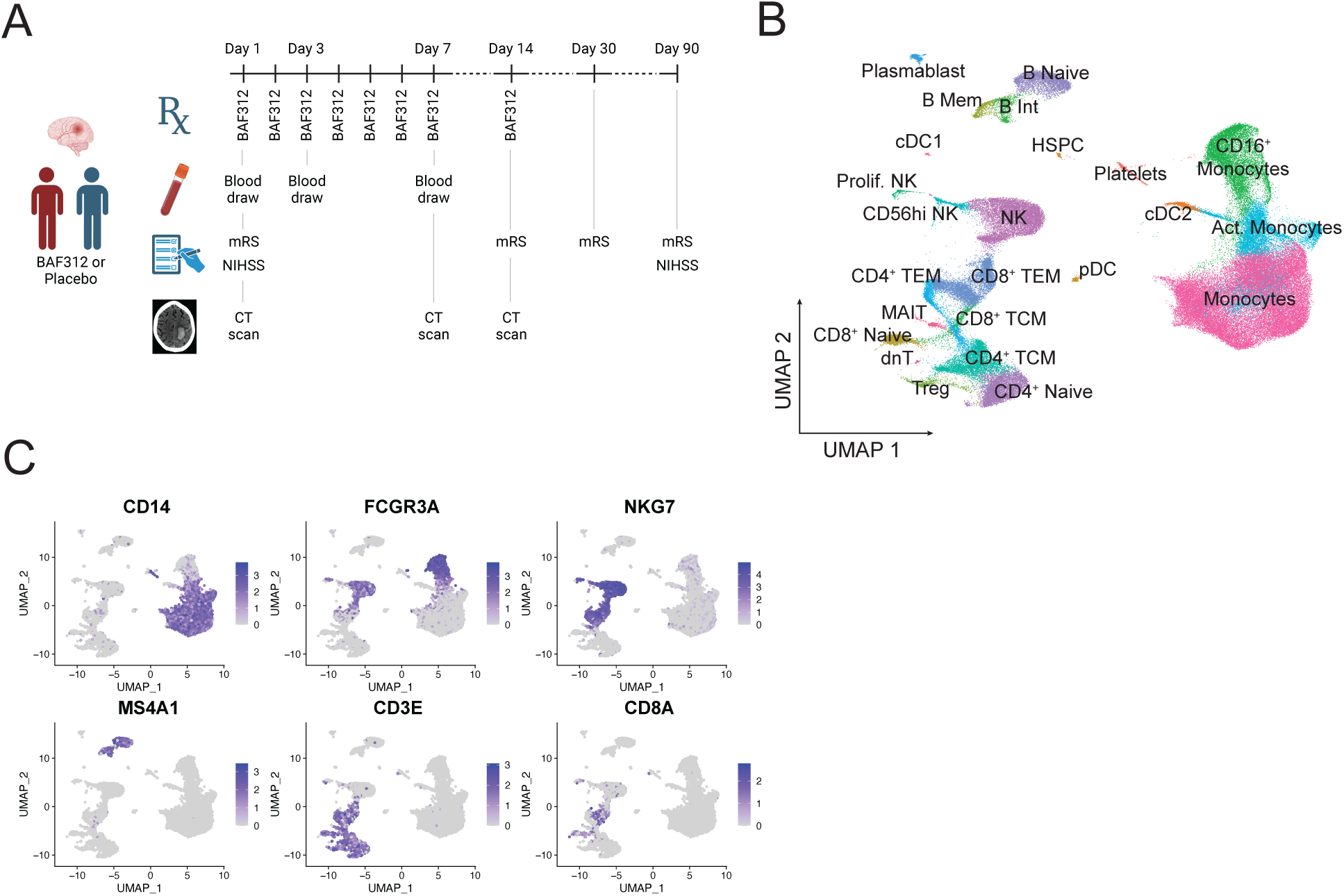
Single cell RNA sequencing analysis pipeline and population identification. (A) Schematic of the clinical trial showing days of treatment with BAF312 or placebo, blood draws, functional tests, and CT scans. (B) UMAP of all 103,301 cells analyzed from the 21 integrated samples after QC filtering. (C) Feature plots showing expression of canonical markers of the lineages analyzed further in the study.

**Table 1.** Patient baseline characteristics. *Samples from this patient were not analyzed by RNA sequencing. **BAF312 treatment in this patient discontinued within 48 hours due to bradycardia. GCS: Glascow Coma Scale. NIHSS: National Institutes of Health Stroke Scale. mRS: Modified Rankin Score.

| Age range<br>(yr) | Sex | ICH Volume<br>(mL) | ICH Location | GCS | NIHSS<br>(Day 1) | mRS<br>(Pre-morbid) | ICH<br>score | Treatment |
| --- | --- | --- | --- | --- | --- | --- | --- | --- |
| 60-69 | F | 14.49 | Basal ganglia | 13 | 18 | 0 | 0 | Placebo* |
| 40-49 | F | 7.28 | Basal ganglia | 14 | 14 | 0 | 0 | BAF312** |
| 40-49 | F | 21.37 | Basal ganglia | 14 | 16 | 0 | 0 | BAF312 |
| 70-79 | F | 45.90 | Basal ganglia | 11 | 19 | 0 | 2 | BAF312 |
| 70-79 | F | 34.32 | Occipital lobe | 15 | 3 | 0 | 1 | Placebo |
| 60-69 | F | 24.48 | Basal ganglia | 12 | 10 | 0 | 1 | Placebo |
| 80-89 | F | 48.47 | Occipital lobe | 15 | 8 | 1 | 2 | BAF312 |
| 60-69 | M | 26.63 | Occipital lobe | 15 | 4 | 0 | 0 | BAF312 |

\* Samples from this patient were not analyzed by RNA sequencing
\*\* BAF312 treatment discontinued within 48 hours due to bradycardia

### Cell population subclustering and pooling into lineages

Patient PBMC were analyzed by single cell RNA-sequencing as described in Methods. Monocytes, CD4^+^ T cells, CD8^+^ T cells, MAIT cells, NK cells, B cells, dendritic cells, HSPCs, and platelets were identified (Fig 1B). Naïve, memory, and activated cells were distinguishable within these lineages (Fig 1B and Supp 1B). For subsequent analyses, cell subsets were pooled into lineages and analysis was focused on the lineages present in high numbers: classical monocytes, CD16^+^ monocytes, NK cells, CD4^+^ T cells, CD8^+^ T cells, and B cells. The identity of these cell lineages is demonstrated by expression of canonical lineage markers (Fig 1C). Proportions of the major cell lineages found in each patient at each timepoint are shown in Supp Fig 1C.

### Peripheral inflammatory response peaked 3 days post ICH

PBMC in healthy individuals are comprised of 10-20% monocytes and 70-90% lymphocytes [53]. Measured by scRNA-Seq, day 1 PBMC were comprised of approximately 50% classical monocytes, 10% CD16^+^ monocytes, 15% NK cells, 15% T cells, 3 % B cells, and small numbers of the other populations shown in Fig 1B. (Fig 2A). Flow cytometric analysis of the same samples confirmed these findings (Supp 2A). Ratios of leukocyte lineages remained relatively constant over days 1-7 after ICH, with no statistically significant changes over time (Fig 2A, Supp 2A).

**Figure 2.**
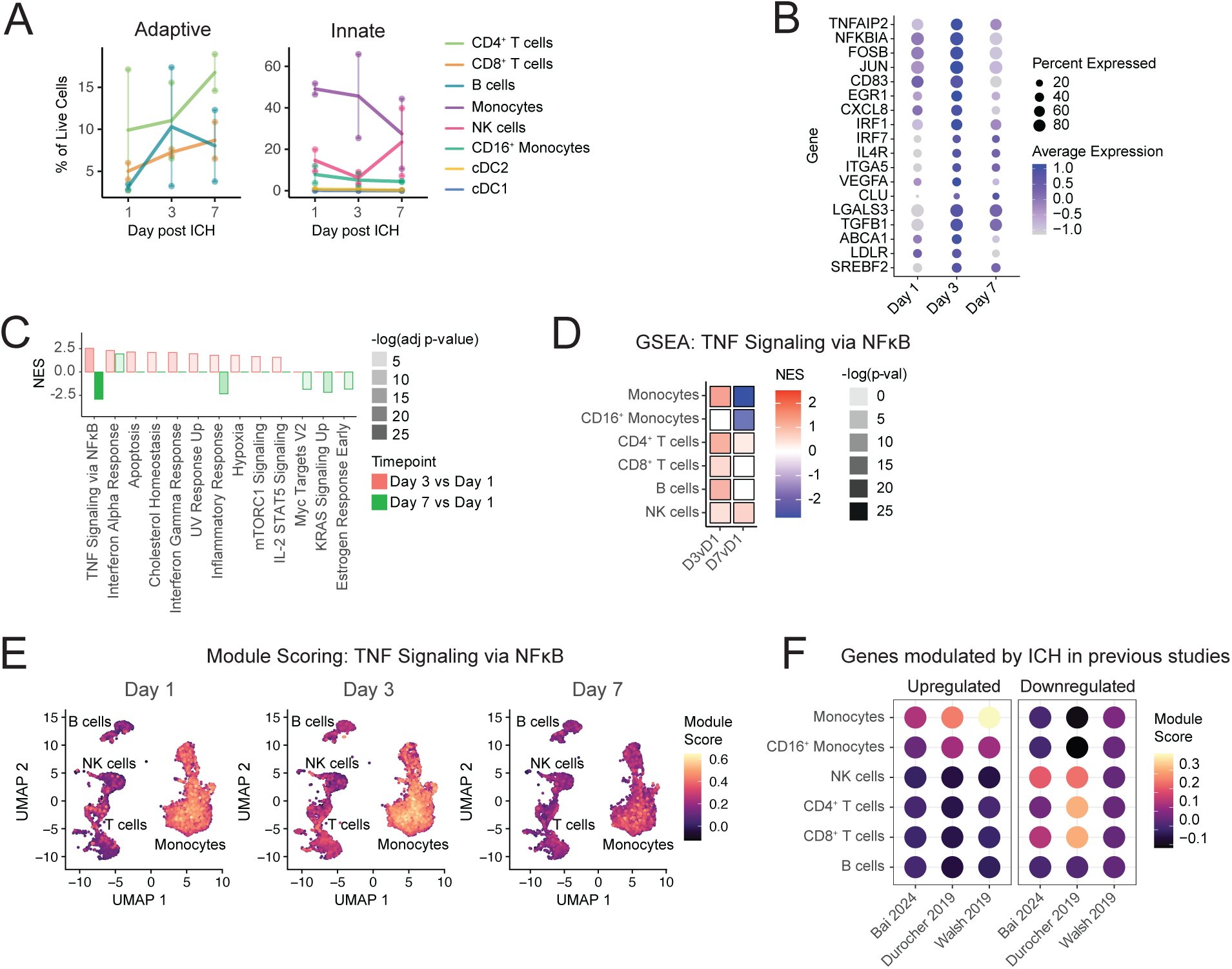
Peripheral inflammatory response peaks 3 days post ICH. (A) Percentages of adaptive (left) and innate (right) immune populations over time after ICH in placebo-treated patients. (B) Time course of select inflammatory gene expression in monocytes from placebo-treated patients after ICH. (C) GSEA with MSigDB Hallmark Pathway genesets of monocytes of placebo-treated patients, Day 3 or Day 7 vs Day 1. (D) GSEA of different cell lineages from placebo-treated patients, comparing Day 3 vs Day 1 and Day 7 vs Day 1, showing only results for MSigDB Hallmark Pathways TNFA_SIGNALING_VIA_NFKB geneset. (E) Module scoring of the TNFA-SIGNALING_VIA_NFKB geneset at days 1, 3, and 7 post ICH. (F) Enrichment at day 1 in our cohort of gene modules modulated by ICH in previous studies. Left panel indicates expression, in our cohort, of genes upregulated in the previous study. Right panel indicates expression, in our cohort, of genes downregulated in the previous study.

Monocytes were quickly activated, upregulating expression of multiple inflammatory response molecules from day 1 to day 3: *FOSB* (LFC 0.35), *JUN* (LFC 0.85), *NFKBIA* (LFC 0.77), *IRF1* (LFC 0.35), and *CXCL8* (LFC 1.1) from day 1 to day 3 after ICH (Fig 2B, Supp Table 3). Expression of these genes peaked at day 3 and decreased from day 3 to day 7: FOSB (LFC -0.63), JUN (LFC -1.1), NFKBIA (LFC -1.5), IRF1 (LFC -2.3), and CXCL8 (LFC -2.8). Geneset enrichment analysis (GSEA) revealed upregulation of multiple inflammatory pathways from day 1 to day 3, including responses to TNF (NES 2.5), IFNα (NES 2.3), and IFNγ (NES 2.1) (Fig 2C, Supp Table 4). The inflammatory response peaked around day 3, as can be seen the change from day 1 to day 7: TNF Signaling via NFκB (NES -2.9), Inflammatory Response (NES -2.3) IFNγ Response (NES not signif.). The IFNα Response remained elevated at day 7 (NES 1.9). The TNF signaling pathway was modulated by all cell populations in our dataset and we focused our downstream analysis on TNF signaling. TNF signaling was increased from day 1 to day 3 for classical monocytes (NES 2.5), CD4^+^ T cells (NES 2.0), CD8^+^ T cells (NES 2.0), B cells (NES 2.4), and NK cells (NES 1.9) (Fig 2D, Supp Table 4). TNF signaling peaked at day 3 for most populations, as can be seen by the change from day 1 to day 7: classical monocytes (NES -2.9), CD16^+^ monocytes (NES -2.7), CD4^+^ T cells (NES 1.7), CD8^+^ T cells (NES not signif.), and B cells (NES not signif.). NK cells were unique among the cell lineages examined in that their TNF signaling signature was more enriched at day 7 vs day 1 (NES 2.2) than day 3 vs day 1 (NES 1.9). Module scoring for the genes present in the MSigDB HALLMARK_TNFA_SIGNALING_VIA_NFKB geneset revealed that this pathway is already highly expressed at day 1 post ICH (Fig 2E). It is more highly upregulated in classical and non-classical monocytes than in other lineages and it is uniformly expressed, not restricted to a subset of classical monocytes (Fig 2E).

Module scoring was performed to examine expression in our dataset of genes that were found to be up- or down-regulated in previous studies: within 48h post-ICH in humans [37], 4-120h post-ICH in humans [38], or 6h post-ICH in swine [39]. Enrichment for these genes in our dataset was found to be similar for cells isolated 1, 3, or 7 days post ICH; results for cells isolated at day 1 are shown. This module scoring revealed that genes previously found by bulk RNA-Seq to be upregulated in total PBMC during ICH in humans and swine [37-39] were enriched in classical monocytes (module score 0.07 to 0.3) and were not enriched in lymphocytes (module score -0.1 to -0.05) in our cohort (Fig 2F). Genes found to be downregulated in human PBMC [37, 38] were enriched in T cells (module score 0.003 to 0.25) and NK cells (module score 0.14 to 0.17) but were not enriched in classical monocytes (module score -0.14 to -0.05) in our cohort (Fig 2F).

### Peripheral inflammatory response decreased by day 3 of ICH in patients treated with BAF312

BAF312 resulted in significant lymphopenia by day 3 of treatment, which was the first timepoint examined post-treatment (Fig 3A). Decreases from day 1 to day 3 were seen in proportions of peripheral CD4^+^ T cells (24% to 1.8%), CD8^+^ T cells (13% to 2.7%), and B cells (6.6% to 2.9%), while an increase was seen in the proportion of monocytes (24% to 61%). These results were confirmed by flow cytometry (Supp 2B). Analysis of lymphocyte subpopulations revealed low day 3 / day 1 ratios of CD4^+^ T_Nv_ (0.024), CD4^+^ T_CM_ (0.074), CD4^+^ T_EM_ (0.14), CD8^+^ T_Nv_ (0.049), CD8^+^ T_CM_ (0.18), and B_Nv_ (0.27) cells in BAF312-treated patients, indicating these populations were reduced in the periphery (Fig 3B). Some lymphocyte populations were not yet reduced at day 3: CD8^+^ T_EM_ (0.91), B_Int_ (1.15), and B_Mem_ (0.94). These populations were decreased later, however, with low day 7 / day 1 ratios: CD8^+^ T_EM_ (0.45), B_Int_ (0.43), and B_Mem_ (0.18). Regulatory T cells were decreased by half, with a day 3 / day 1 ratio of 0.47 and a day 7 / day 1 ratio of 0.54.

**Figure 3.**
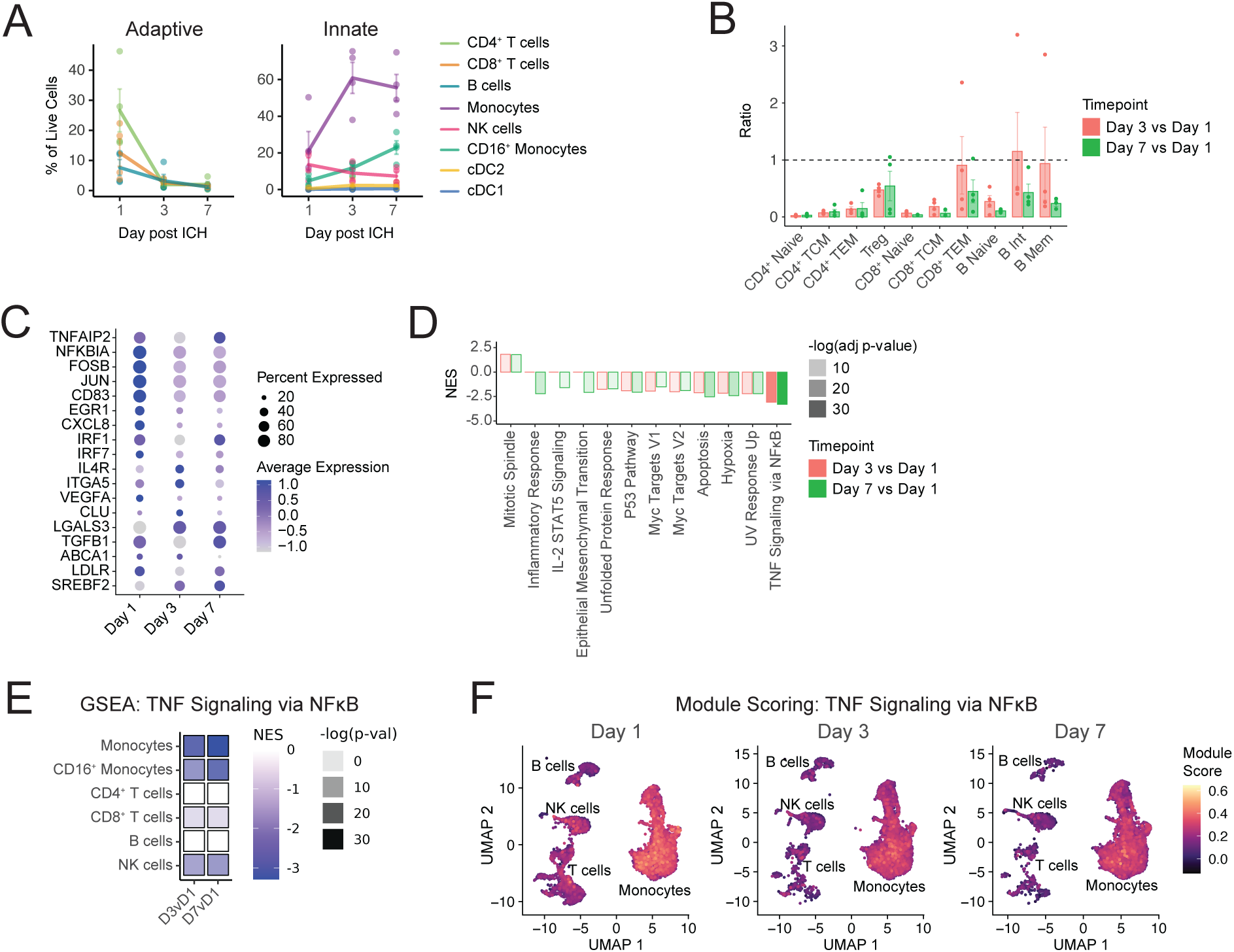
Peripheral inflammatory response decreases by day 3 of ICH in patients treated with BAF312. (A) Percentages of adaptive (left) and innate (right) immune populations over time after ICH in BAF312-treated patients. (B) Ratios of lymphocyte subpopulations in BAF312-treated patients at Day 3 or Day 7 vs Day 1. (C) Time course of select inflammatory gene expression in monocytes from BAF312-treated patients after ICH. (D) GSEA with MSigDB Hallmark Pathway genesets of monocytes of BAF312-treated patients, Day 3 or Day 7 vs Day 1. (E) GSEA of different cell lineages from BAF312-treated patients, comparing Day 3 vs Day 1 and Day 7 vs Day 1, showing only results for MSigDB Hallmark Pathways TNFA_SIGNALING_VIA_NFKB pathway. (F) Module scoring of the TNFA-SIGNALING_VIA_NFKB geneset at days 1, 3, and 7 post ICH.

In contrast to what was seen in placebo-treated patients, inflammatory gene expression by monocytes in BAF312-treated patients was generally downregulated from day 1 to day 3: *FOSB* (LFC -0.70), *JUN* (LFC -0.74), *NFKBIA* (LFC -1.01), *IRF1* (LFC -0.32), and *CXCL8* (LFC -1.91) (Fig 3C, Supp Table 5). Expression of these genes stayed relatively constant from day 3 to day 7: *FOSB* (LFC 0.087), *JUN* (LFC not signif.), *NFKBIA* (LFC not signif.), and *CXCL8* (LFC not signif.). *IRF1* was an exception, increasing from day 3 to day 7 (LFC 0.40). GSEA found decreases in TNF Signaling via NFκB pathway genes (NES -3.1) from day 1 to day 3 and in IL-2 STAT5 Signaling (NES -1.6) and Inflammatory Response pathway genes (NES -2.2) from day 1 to day 7 post ICH (Fig 3D and Supp Table 6). Genes in the TNF Signaling via NFκB geneset also decreased from day 1 to day 3 in CD16^+^ monocytes (NES -2.9), CD8^+^ T cells (NES -2.1), and NK cells (NES -2.7) in BAF312-treated patients (Fig 3E, Supp Table 6). Module scoring indicated that genes in the TNF signaling via NFκB geneset decreased uniformly within each lineage, suggesting that BAF312 did not target specific subsets of these lineages (Fig 3F).

### BAF312 decreased inflammatory response compared to placebo

Monocytes from patients treated with BAF312, compared with placebo, had lower expression of canonical inflammation-associated genes, including *NKFBIA* (LFC -1.8), *FOSB* (LFC -0.86), *JUN* (LFC -1.4), and *CXCL8* (LFC -4.2) at day 3 post ICH (Fig 4A, Supp Table 7). Similar effects were seen, though at lower magnitude, at day 7: *NKFBIA* (LFC -0.37), *FOSB* (LFC not signif.), *JUN* (LFC -0.26), and *CXCL8* (LFC -1.4). GSEA revealed that at day 3, BAF312 treatment was associated with decreased monocyte expression of genes associated with responses to TNF (NES -3.2), IFNα (NES -1.8), IFNγ (NES -2.1), hypoxia (NES -2.1), and general inflammation (NES -2.3) (Fig 4B, Supp Table 8). GSEA found no impact of BAF312 on these pathways at day 7. The suppressive effect of BAF312 on the day 3 response to TNF Signaling via NFκB was also seen in CD16^+^ monocytes (NES -2.8), CD4^+^ T cells (NES -2.3), CD8^+^ T cells (NES -1.9), and B cells (NES -2.0) (Fig 4C, Supp Table 8). BAF312 did not impact the TNF Signaling pathway in NK cells at day 3. At day 7, BAF312 decreased TNF Signaling in NK cells (NES -1.9), CD4^+^ T cells (NES -2.2), and B cells (NES -1.7).

**Figure 4.**
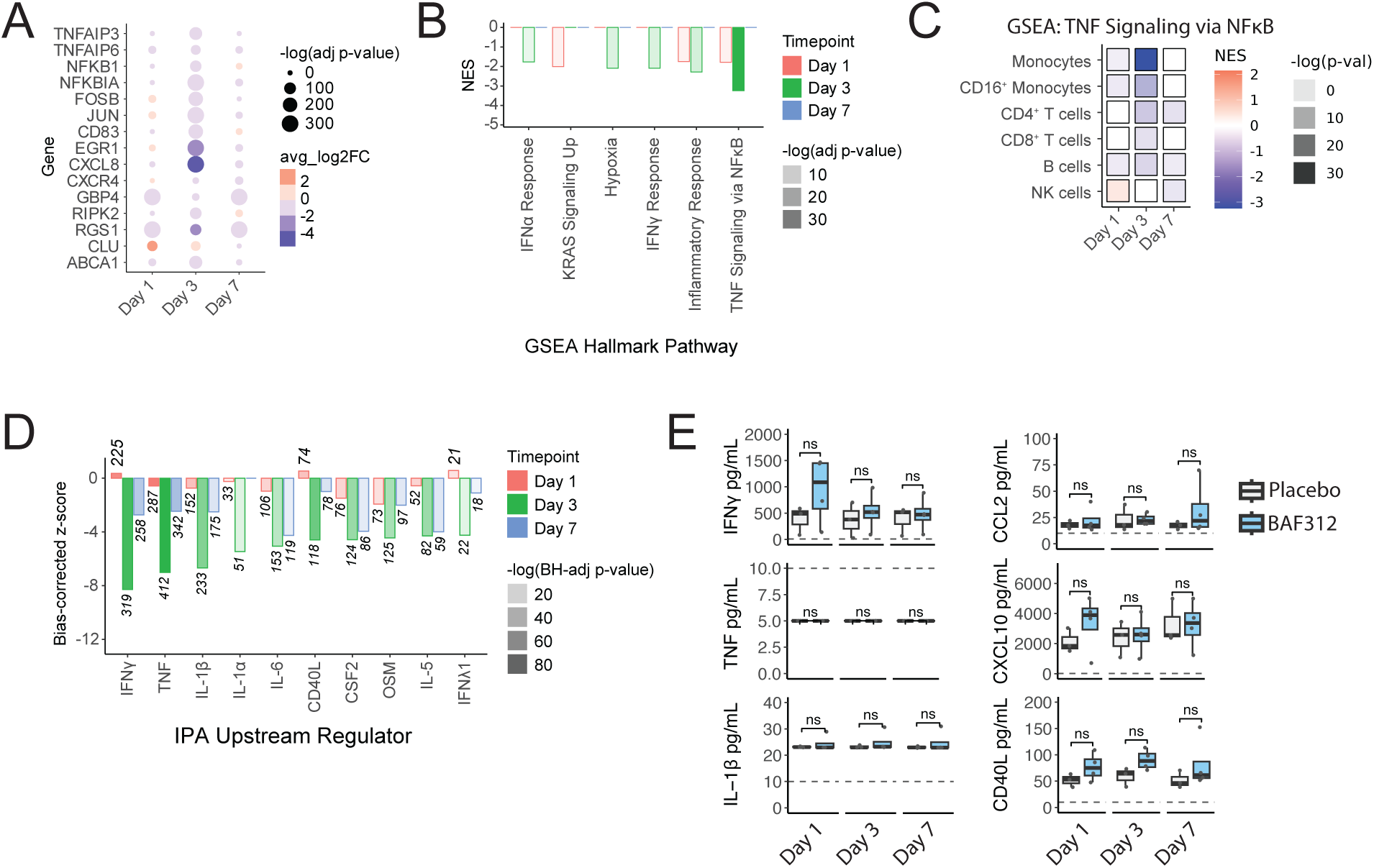
BAF312 decreased inflammatory response compared to placebo. (A) Differential expression of select inflammatory genes in BAF312-treated vs placebo-treated patients. (B) GSEA of monocytes from BAF312-treated vs placebo-treated patients. (C) GSEA of different cell lineages in BAF312-treated vs placebo-treated patients, showing only results for MSigDB Hallmark Pathways TNFA_SIGNALING_VIA_NFKB pathway. (D) Enrichment of cytokine signaling pathways in BAF312-treated vs placebo-treated patients, measured by Ingenuity Pathways Analysis Upstream Regulators Analysis. Numbers on bars indicate the number of genes overlapping with the pathway. (E) Concentration of cytokines in patient plasma, measured by Cytometric Bead Array. Dashed lines indicate lower limit of detection.

Among the 7 patients whose samples were analyzed by RNA sequencing, ICH score was similar between the placebo-treated (average ICH score: 1) and BAF312-treated (average ICH score: 0.8) groups (Table 1). However, differences in initial stroke severity nonetheless had the potential to confound the direct comparison of patients who received the placebo to those who received BAF312 treatment. Transcript counts were therefore pseudobulked and ICH score was incorporated into the linear model used to calculate differential gene expression. The statistical significance of identified DEGs was lost after pseudobulking, but the trend of BAF312 reducing inflammatory gene expression was maintained (Supp 3A-C).

Ingenuity Pathway Analysis (IPA) Upstream Regulators Analysis was used to identify cytokines that could be responsible for the decrease in inflammatory response seen in BAF312-treated patients. This revealed that patients receiving BAF312 treatment had decreased expression of signaling pathways for multiple inflammatory cytokines, including IFNγ (z-score -8.2), TNF (z-score -7.0), IL-1β (z-score -6.7), and IL-6 (z-score -5.1) at day 3 post ICH (Fig 4D, Supp 3D). This impact persists, though at a lower magnitude, at day 7: IFNγ (z-score -2.7), TNF (z-score -2.5), IL-1β (z-score -2.5), and IL-6 (z-score -4.3) Cytokine and chemokine concentrations were assessed in plasma from these patients by cytokine bead array (BD Biosciences) and revealed no significant differences in the concentrations of IFNγ, TNF, IL-1β, CCL2, CXCL10, CD40L, IL-1α, IL-6, IL-4, or IL-10 (Fig 4E, Supp 3E).

### Increased TNF response after ICH was associated with better functional outcome

The correlation of disease severity at presentation with the monocyte response was then examined. Patients were bifurcated into groups based on clinical measures of ICH severity and outcome and GSEA was performed to identify transcriptional pathways that differed between these groups. GSEA revealed that higher day 1 hematoma volume (> 30mL) was associated with higher TNF signaling at day 3 (NES 2.2) (Fig 5A). Greater day 1 perihematomal edema volume (> 30mL) was also associated with higher TNF signaling at day 3 (NES 2.3) (Fig 5B). Higher relative perihematomal edema (> 1) at day 1 was associated with increased TNF signaling for the entire first week post ICH (day 1 NES 2.3, day 3 NES 2.6, day 7 NES 2.2) (Fig 5C). Surprisingly, higher day 1 NIH Stroke Scale (NIHSS) score (> 9), indicating a more severe stroke, correlated with less TNF signaling at day 1 (NES -1.9) and day 3 (NES -1.9) post ICH (Fig 5D).

**Figure 5.**
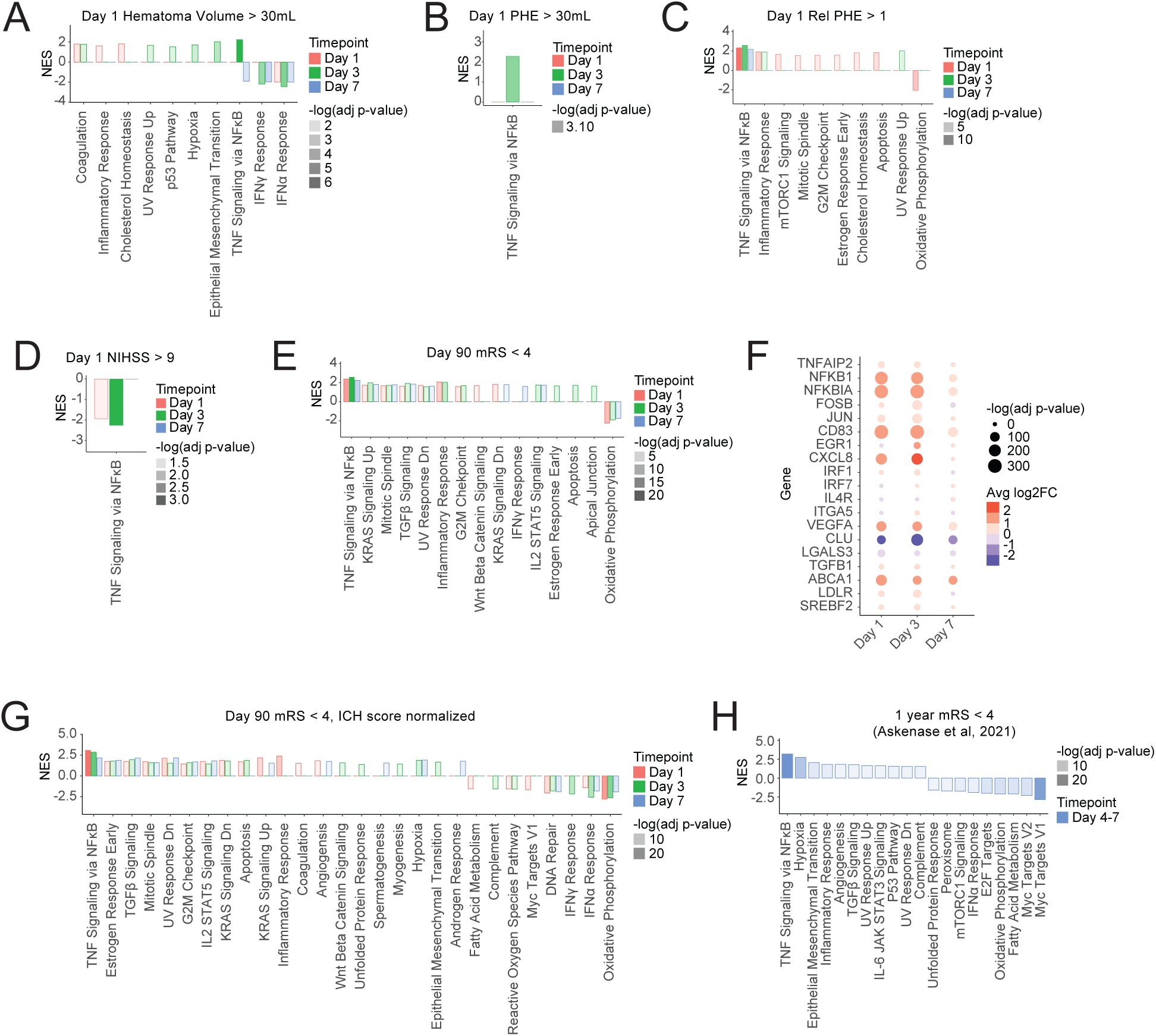
Increased TNF response was associated with better functional outcome. (A-E) GSEA of monocytes, comparing patients with: Day 1 hematoma volume greater than 30mL vs less-than 30mL (A); Day 1 perihematomal edema greater than 30mL vs less-than 30mL (B); Day 1 relative perihematomal edema greater than 0.5 vs less than 0.5 (C); Day 1 NIHSS of greater-than 9 vs 1-9 (D); Day 90 mRS of 0-3 vs 4-6 (E). (F) Differential expression of select inflammatory genes in patients with Day 90 mRS of 0-3 vs 4-6. (G) GSEA as in panel E, but after downsampling and fitting to a linear model that normalizes for ICH score. (H) GSEA of monocytes/macrophages isolated from the hematoma of ICH patients, comparing mRS of 0-3 vs 4-6 at 1 year post ICH, from Askenase, et al 2021.

Patients with better functional outcome (Modified Rankin Score, mRS, below 4) at day 90 were found to have monocytes with greater TNF signaling throughout the first week after ICH (day 1 NES 2.4, day 3 NES 2.6, day 7 NES 2.2) (Fig 5E). These patients also displayed less oxidative phosphorylation in the first week of ICH (day 1 NES -2.2, day 3 NES -1.9, day 7 NES -1.7) (Fig 5E). Individual gene expression revealed that lower day 90 mRS was associated with higher expression at day 1 and day 3 of a subset of the inflammatory genes that are upregulated during ICH, notably *NFKB1* (day 3 LFC 1.1), *NFKBIA* (day 3 LFC 1.3), *CXCL8* (day 3 LFC 3.0), and *VEGFA* (day 3 LFC 1.2) (Fig 5F, 2B). Full results, for all cell types, of the gene expression and GSEA pathways that correlate with various patient characteristics can be found in Supp Tables 9 and 10. To control for variation in initial ICH severity, samples were pseudobulked and gene expression was fit to a linear model in which ICH score was included. Controlling for initial ICH severity did not change the correlation of lower day 90 mRS with increased TNF signaling and decreased oxidative phosphorylation (Fig 5G).

To confirm these results, a similar analysis was performed on a previously published dataset reporting transcriptional responses of monocytes isolated from patients participating in the MISTIE III trial [35]. We found that in the MISTIE III dataset, in monocytes isolated from the peripheral blood, good outcome was associated with lower oxidative phosphorylation (NES -2.2), but also lower TNF signaling (NES -2.3) (Supp 4A). We also found that monocytes isolated from hematomas of patients with good outcome (mRS < 4) in the MISTIE III trial had increased TNF signaling (NES 3.2) and decreased oxidative phosphorylation (NES -2.1), similar to the findings in the peripheral blood in our cohort (Fig 5H).

## DISCUSSION

The longitudinal samples collected in this study enabled transcriptome-wide characterization of the peripheral immune response in the first week after intracerebral hemorrhage. While the number of patients in the study was small, this is the first comprehensive study of the effect of S1P receptor modulation in patients with acute ICH, with robust clinical data and outcomes measured throughout the trial and reported in the supplemental tables of this study. This study characterized the responses of classical and non-classical monocytes, CD4^+^ T cells, CD8^+^ T cells, B cells and NK cells. Patients from both the placebo and treatment arm demonstrated moderate lymphopenia at day 1, which has been previously reported in ICH and is associated with larger hemorrhage volume and decreased survival [54, 55]. This lymphopenia was dramatically augmented at days 3 and 7 in patients receiving BAF312, confirming that BAF312 had a robust effect within this subacute stage of ICH. A previous report found an increase in peripheral blood monocytes at days 3-5 post ICH [56]. The present study found no statistically significant changes in monocyte numbers after ICH, possibly due to the small number of patients recruited in this study.

The inflammatory response was found to peak 3 days post ICH in placebo-treated patients, in agreement with previous reports [35, 56]. The contribution of TNF to pathological neuroinflammation during ICH is well documented [7, 35, 43, 57] and the MSigDB Hallmark TNF signaling via NFκB geneset was upregulated post ICH in all of the cell lineages examined in this study. This geneset was used throughout the study as a readout of the inflammatory response. Monocytes expressed this geneset at high levels even by day 1 after ICH. This complements previous findings that TNF is produced in the rat brain as early as 2 hours post ICH [43]. Previous reports found that expression of type I inflammation-associated molecules in the hematoma and CSF are highest in the first 1-4 days after ICH and decrease by day 7 [7, 35, 57]. The findings in the present study suggest that the immune responses in the peripheral blood resemble those in the hematoma. The response of most major blood leukocyte lineages peaked at day 3 post-ICH, which agrees with previous studies that have found that the immune response becomes less inflammatory and more reparative after day 3 post-ICH [7, 35]. NK cells were the notable exception, remaining at peak activation through day 7.

Module scoring revealed that genes shown to be upregulated during ICH in previous studies published by Walsh et al. [39], Bai et al. [37], and Durocher et al. [38] were highly expressed in our cohort as well. This demonstrates the consistency and reliability of these studies and provides additional insights possible with single cell RNA-Seq. The previous studies used bulk-RNA-Seq to analyze unsorted PBMC or whole peripheral blood. We found that the gene signatures upregulated in these studies were predominantly expressed by monocytes in our cohort, with minimal expression in other populations. This suggests that the monocyte response dominates the peripheral blood transcriptional response during the acute stage of ICH. Additionally, genes downregulated in bulk PBMC after ICH were highly expressed by T cells and NK cells in our cohort, suggesting a decrease in numbers or activation levels of these populations relative to monocytes.

A subset of the patients in this study was treated with the immunomodulatory drug BAF312. The peripheral immune response was found to be profoundly suppressed in these patients. The frequencies of peripheral blood CD4^+^ T cells, CD8^+^ T cells, and B cells were significantly decreased by day 3 and remained decreased at day 7 post ICH. Naïve and central memory populations were the first to decrease, but by day 7 the memory cells were significantly reduced as well. This complements the finding that effector T cells and transitional B cells are resistant to reduction from circulation by BAF312 after 6-12 months of treatment in patients with multiple sclerosis [12, 15]. The innate immune response was also affected, with the inflammatory signature of monocytes from BAF312-treated patients decreasing at days 3 and 7 post ICH.

The study discussed monocytes in the most depth due to the established importance of monocytes in the first week of ICH, the relative paucity of information about the impacts of BAF312 on monocytes, and because analysis of lymphocytes in BAF312-treated patients was complicated by the reduction of peripheral lymphocyte numbers by BAF312. BAF312 treatment was associated with significantly inhibited inflammatory gene expression in monocytes at day 3 post ICH. Inflammatory gene expression stayed low through day 7, by which point monocytes in placebo-treated patients had also decreased their inflammatory gene expression. BAF312 therefore appears to have accelerated the resolution of the inflammatory response of peripheral blood monocytes. Ingenuity Pathways Analysis revealed that BAF312 also suppressed expression of signaling pathway genes for IFNγ, IL-1β, CD40L, and other inflammatory genes. This led to the hypothesis that BAF312 treatment reduced the concentration of these cytokines in the blood. Measurement of cytokine concentration in plasma samples from these patients did not reveal an effect of BAF312. This agrees with previous studies, finding that FTY720 does not decrease serum inflammatory cytokine concentration in humans or during murine MCAO [32, 58]. It is possible that BAF312 limits the ability of monocytes to respond to the cytokines present in their environment.

The correlation of ICH severity with peripheral inflammation was also examined. Greater volume of hematoma and perihematomal edema were correlated with increased TNF signaling in monocytes at day 3. Relative perihematomal edema had an even stronger association with inflammation, correlating with increased TNF signaling for the entire first week post ICH. These results are consistent with previous studies finding that CCL2, IL-1β, and MMPs promote edema [59]. In contrast, a recent cohort study of patients with BBB permeability quantified by dynamic contrast-enhanced MRI at a median of 56 hours after ICH found no correlation between microglial activation and edema volume. That study also found an inverse correlation between BBB permeability and edema volume, calling into question the roles of inflammation and BBB permeability in driving edema during this time frame after ICH [60].

The correlation of acute-stage monocyte gene expression with long-term functional outcome was also examined. This has the potential to reveal biological pathways that influence functional outcome, as well as to identify early biomarkers of long-term outcome. Good mRS (mRS < 4) at day 90 was highly correlated with gene expression in the first week of ICH. Indeed, in this cohort, the number of pathways identified, their normalized enrichment scores, and their statistical significance were greater for correlations of gene expression with mRS than with hematoma volume, perihematomal edema, or day 1 NIHSS. The patients with good day 90 mRS had, surprisingly, higher TNF signaling throughout the first week after ICH. We also found that increased TNF signaling was associated with greater relative perihematomal edema at day 1. The correlation of TNF signaling with good outcome appears to be in contrast with studies finding that blockade or genetic ablation of TNF improves behavioral outcome in murine models of ICH [43-46]. This apparent paradox may be explained by the finding that infiltrating macrophages contribute to breakdown of hematoma and resolution of disease in the subacute stage of ICH [7]. It is possible that the beneficial effects of infiltrating macrophages and other TNF-producing cells outweigh the direct neurotoxic effects of TNF during ICH. Indeed, increased microglial activation has been associated with improved day 90 mRS [60]. Additionally, TNF has neuroprotective effects during ischemic stroke that may also be relevant during ICH [61, 62]. Larger studies will be able to examine in greater resolution the immune response during ICH, in particular the pro-inflammatory and reparative roles that TNF-producing macrophages play and the connection between TNF and perihematomal edema.

## LIMITATIONS

This study is limited by multiple factors. Samples were from a clinical trial that was prematurely terminated, which reduced the number of samples that could be collected and limited the statistical power of our results. This also resulted in underrepresentation of male patients in our substudy, precluding analysis of sex effects in our dataset. Additionally, the samples were shipped overnight before analysis, during which time there might have been changes in gene expression or plasma cytokine concentration. Furthermore, confirmation of the RNA-sequencing results was limited to flow cytometry, measurement of plasma cytokine concentrations, and comparisons with previous studies. The impact of BAF312 on TNF signaling and the role of TNF in ICH were not directly confirmed by experiments.

## Supporting information

Supp Table 1 - Cellranger Sequencing Metrics

Supp Table 2 - Demographics and Clinical Characteristics

Supp Table 3 - DEG Placebo by Timepoint

Supp Table 4 - GSEA Placebo by Timepoint

Supp Table 5 - DEG BAF312 by Timepoint

Supp Table 6 - GSEA BAF312 by Timepoint

Supp Table 7 - DEG by Treatment

Supp Table 8 - GSEA by Treatment

Supp Table 9 - DEG by Clinical Characteristics

Supp Table 10 - GSEA by Clinical Characteristics

Supp Table 11 - Supporting Data Values

## NON-STANDARD ABBREVIATIONS

BBB: blood-brain barrier
CBA: cytometric bead array
GSEA: gene set enrichment analysis
ICH: intracerebral hemorrhage
LFC: log-fold change
mRS: modified Rankin score
NES: Normalized enrichment score
NIHSS: National Institutes of Health Stroke Scale
S1P: sphingosine-1-phosphate
S1PR: sphingosine-1-phosphate receptor

## AUTHOR CONTRIBUTIONS

JHD: designing research studies, conducting experiments, acquiring data, analyzing data, writing the manuscript; SD: analyzing data, editing the manuscript; KS, JHC, CJM, PGMW, NP: acquiring data; AH, HEH, KBW: acquiring data, editing the manuscript; LHS: designing research studies, analyzing data, writing the manuscript

## ACKNOWLEDGEMENTS

Research reported in this publication was supported by NIH NHLBI Vascular Research Training Grant Number T32HL007950 (JHD), AHA Postdoctoral Fellowship Number 830877 (JHD), AHA Established Investigator Award Number 19EIA34770133 (LHS).

Novartis supported the BAF312 clinical trial but provided no financial support or analyses for this substudy.

We thank the Yale Flow Cytometry Core for their assistance. They are supported in part by an NCI Cancer Center Support Grant # NIH P30 CA016359.

We thank the Yale Center for Genome Analysis for their assistance. They are supported in part by the National Institute of General Medical Sciences of the National Institutes of Health Award Number 1S10OD030363-01A1.

We thank the study coordinators who organized the collection and shipping of samples. We thank the patients who took part in this clinical trial.

## FIGURE LEGENDS

**Supp Fig 1.**
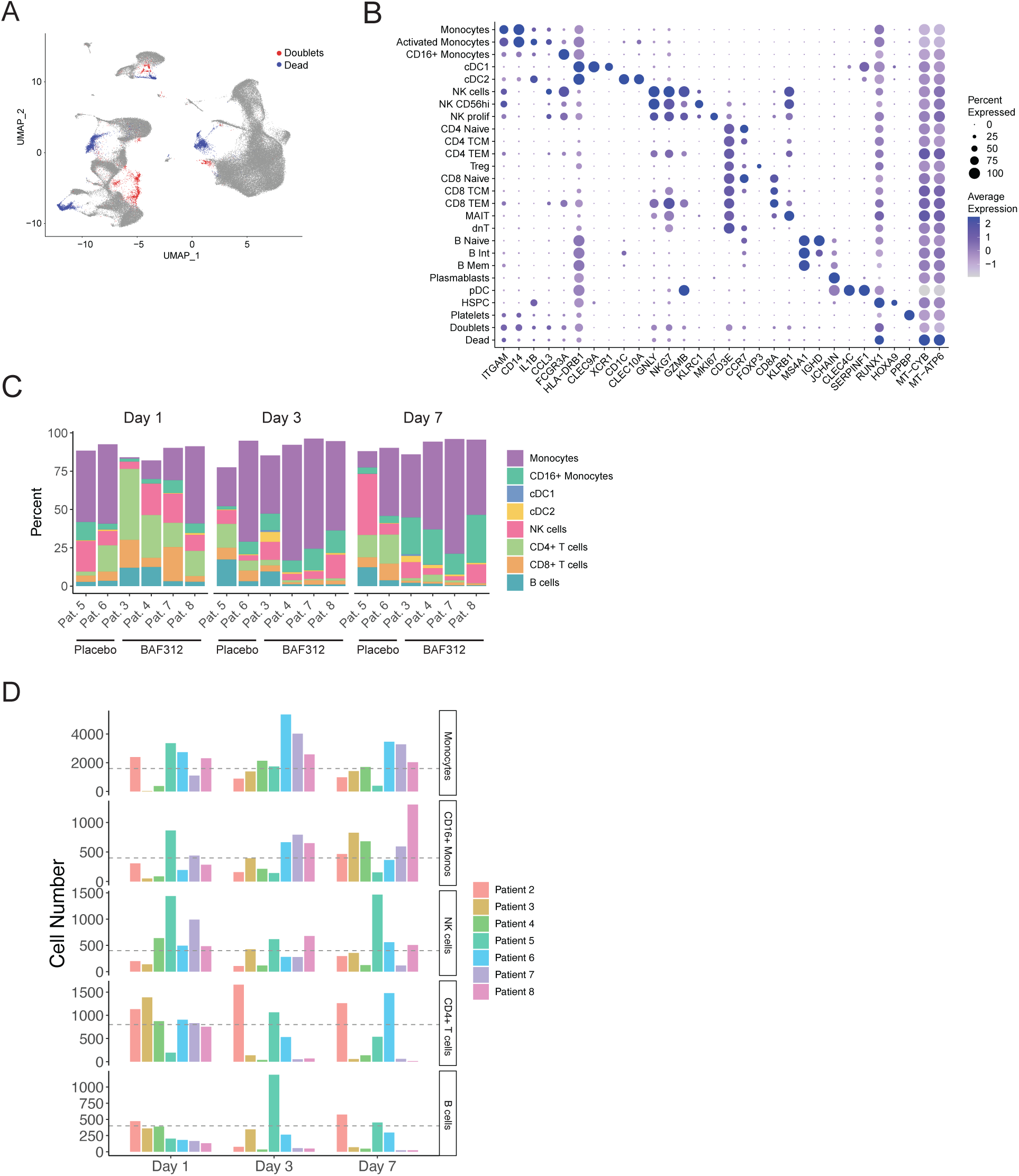
Clustering and downsampling. (A) UMAP including doublets and dead cells that were removed before downstream analysis. (B) Expression dotplot of select genes that help identify the indicated subclusters of cells. (C) Proportions of cells found in select major cell lineages, in each patient, at each timepoint. Proportions do not add to 100% because minor lineages are not represented. (D) Cell numbers collected per lineage, per sample. Dashed lines indicate the number of cells to which each cell type was downsampled.

**Supp Fig 2.**
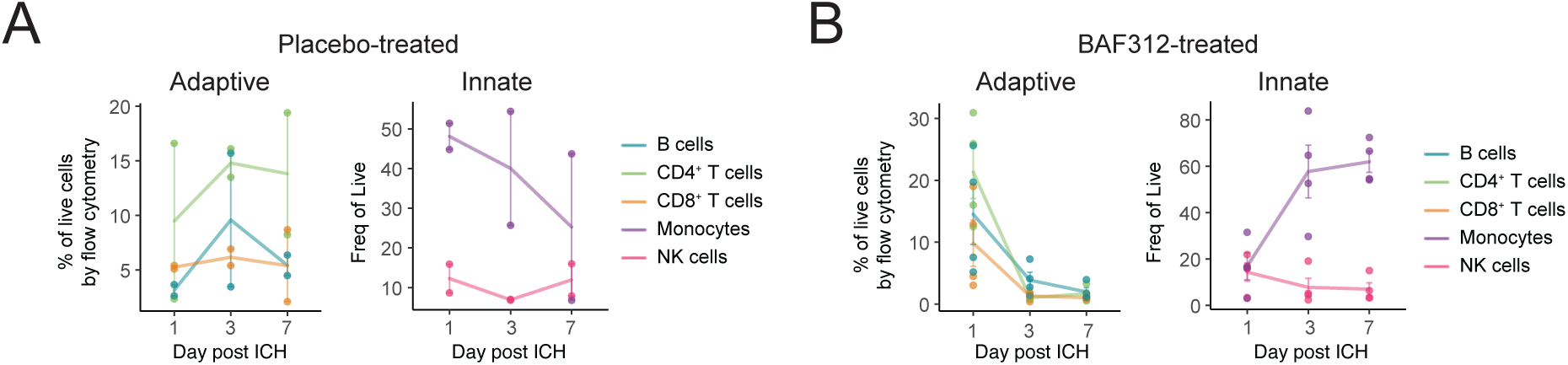
Population frequency changes after ICH by flow cytometry. (A-B) Cell proportions in placebo-treated patients (A) and BAF312-treated patients (B), measured by flow cytometry. Adaptive immune populations are shown on the left and innate immune populations are shown on the right.

**Supp Fig 3.**
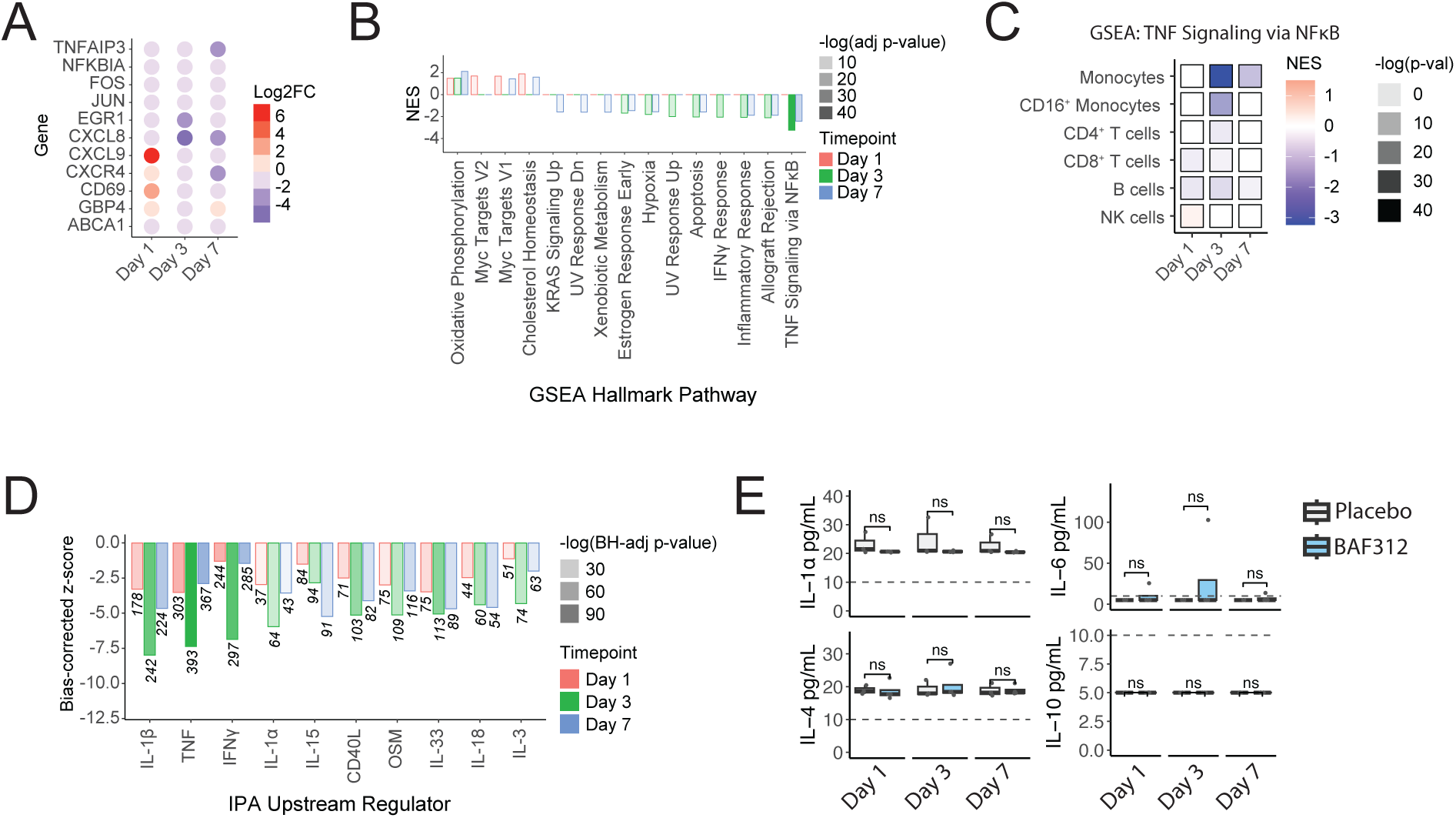
BAF312 decreased inflammatory response compared to placebo. (A-D) Samples were pseudobulked by cell type and fit to a linear model in which gene expression is normalized for ICH score. (A) Differential expression of select inflammatory genes in monocytes from BAF312-treated vs placebo-treated patients. (B) GSEA of monocytes from BAF312-treated vs placebo-treated patients. (C) GSEA of different cell lineages in BAF312-treated vs placebo-treated patients, showing only results for MSigDB Hallmark Pathways TNFA_SIGNALING_VIA_NFKB pathway. (D) Enrichment of cytokine signaling pathways in BAF312-treated vs placebo-treated patients, measured by Ingenuity Pathways Analysis Upstream Regulators Analysis. (E) Concentration of cytokines in patient plasma, measured by Cytometric Bead Array. Dashed lines indicate lower limit of detection.

**Supp Fig 4.**
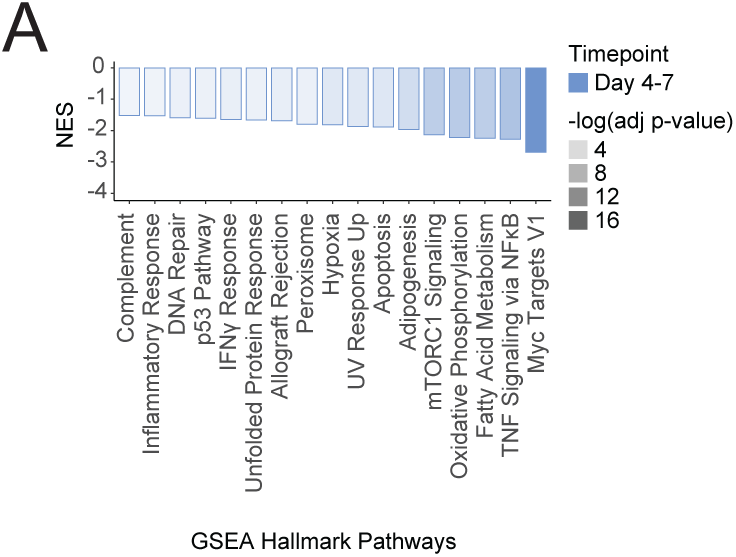
Association of outcome with immune response in MISTIE III study. GSEA of monocytes isolated from the peripheral blood of ICH patients, comparing mRS of 0-3 vs 4-6 at 1 year post ICH, from Askenase, et al 2021.

## SUPPLEMENTAL TABLE LEGENDS

**Supp Table 1. Sequencing metrics. Sequencing quality metrics as reported by Cell Ranger v7.1.0**

**Supp Table 2. Patient demographics and clinical characteristics.** Patient demographics, medical history, central reading data, and clinical outcome measurements.

**Supp Table 3. DEG of Placebo-treated patients – comparisons by timepoint.** Differential gene expression, performed using the FindMarkers() function as described in Methods. DEGs were calculated for each of the 6 main lineages identified. Three comparisons were made for each population in placebo-treated patients: Day 3 vs Day 1, Day 7 vs Day 3, and Day 7 vs Day 1.

**Supp Table 4. GSEA of Placebo-treated patients – comparisons by timepoint.** GSEA was performed using the fgsea() function of the fgsea package in R. Input genes were the DEGs shown in Supp Table 3, ranked by log_2_(fold-change).

**Supp Table 5. DEG of BAF312-treated patients – comparisons by timepoint.** Differential gene expression, performed using the FindMarkers() function as described in Methods. DEGs were calculated for each of the 6 main lineages identified. Three comparisons were made for each population in BAF312-treated patients: Day 3 vs Day 1, Day 7 vs Day 3, and Day 7 vs Day 1.

**Supp Table 6. GSEA of BAF312-treated patients – comparisons by timepoint.** GSEA was performed using the fgsea() function of the fgsea package in R. Input genes were the DEGs shown in Supp Table 5, ranked by log_2_(fold-change).

**Supp Table 7. DEG of placebo-treated vs BAF312-treated patients.** Differential gene expression, performed using the FindMarkers() function as described in Methods. DEGs were calculated for each of the 6 main lineages identified. Three comparisons were made for each population: placebo-treated vs BAF312-treated at Day 1, Day 3, and Day 7.

**Supp Table 8. GSEA of placebo-treated vs BAF312-treated patients.** GSEA was performed using the fgsea() function of the fgsea package in R. Input genes were the DEGs shown in Supp Table 7, ranked by log_2_(fold-change).

**Supp Table 9. DEG of pooled patients by clinical characteristics.** Differential gene expression, performed using the FindMarkers() function as described in Methods. DEGs were calculated for each of the 6 main lineages identified. Patients were bifurcated into groups based on clinical measures of ICH severity and outcome and DEGs were identified that distinguish these groups at Day 1, Day 3, and Day 7.

**Supp Table 10. GSEA of pooled patients by clinical characteristics.** GSEA was performed using the fgsea() function of the fgsea package in R. Input genes were the DEGs shown in Supp Table 9, ranked by log_2_(fold-change).

**Supp Table 11. Supporting Data Values.** Raw data values are included for all plots displaying non-transcriptome data.

